# Development and Validation of a Risk Prediction Model for In-Hospital Mortality of Adult Acute Kidney Injury Patients Requiring Intermittent Hemodialysis, Gondar, Northwest Ethiopia

**DOI:** 10.1101/2025.08.01.25332572

**Authors:** Awoke Worku Dubale, Dessie Abebaw Angaw, Lakew Asmare Tilaye

## Abstract

**Introduction:** Acute kidney injury is a common complication in hospitalized patients and is strongly associated with increased morbidity, mortality, and healthcare costs. In spite of advancements in dialysis techniques and critical care, mortality rates in acute kidney injury patients requiring intermittent hemodialysis remain high, depending on the severity of illness and underlying comorbidities. This Study aimed to develop and validate a risk prediction model of In-hospital mortality for adult acute kidney injury patients requiring intermittent hemodialysis.

**Method:** We retrospectively recruited 829 patients admitted to University of Gondar and Felege Hiwot Comprehensive Specialized Hospitals from 2 October 2016 to 30 March 2025.Demographic, clinical and laboratory candidate variables were selected based on a literature review and expert opinion. A stepwise backward elimination technique was used and the role of each predictor in the multivariable analysis was assessed by the Akaike *Information Criterion*. Presence of multicollinearity was checked using variance inflation factor .Bootstrapping technique was used for internal validation cohorts. We assessed model discrimination using the area under the receiver operator characteristic curve. Clinical utility and applicability of the model was assessed with decision curve analysis and nomogram respectively.

**Result:** A total of 829 adult patients were included in the study. Of these 247 (29.8%) (95%CI: 24.53-32.53) patients died during hospitalization. The predictive model based on seven predictors (age, vasopressor, mechanical ventilation, immunosuppression, cardiovascular disease, platelet count, and serum creatinine) exhibited a predictive performance, as assessed by AUC of 0.85(95%CI: .81-0.89) in development dataset, and 0.8(95%CI: .77-.83) in internal validation for the discrimination of the model.

**Conclusion:** This study successfully developed and internally validated a risk prediction model for in-hospital mortality in adult patients with acute kidney injury requiring intermittent hemodialysis. The model demonstrates good potential to support clinical decision-making by enabling individualized mortality risk estimation at the time of initiating renal replacement therapy. With external validation in independent cohorts, it could become a valuable tool to guide prognosis and facilitate shared decision-making in clinical practice.

## 1. Introduction

Higher rates of morbidity and death during and after hospitalization are associated with acute kidney injury requiring dialysis (AKI-D) [1]. AKI-D incidence increased by 10% every year, according to a nationwide study carried out in the United States [2, 3]. In the outpatient phase after hospital discharge, up to 30% of AKI-D patients may require routine hemodialysis. Mortality rates for patients with AKI-D have been recorded at various time points; rates for in-hospital, 90-day, and 6-month periods were 35%, 45%, and 49%, respectively [4]. Approximately 13 million people suffering from this worldwide and 1.7 million deaths every year and associated with high expenditures in healthcare systems, reaching 1.72 billion dollars per year [5].

Intermittent hemodialysis (IHD) is a popular renal replacement treatment for patients with severe AKI. However, mortality rates for AKI patients receiving IHD remain high, ranging from 30% to 60%, depending on the severity of disease and underlying comorbidities, despite advancements in critical care and dialysis procedures [6].

According to earlier research, scores like the Acute Physiology and Chronic Health Evaluation (APACHE), the Simplified Acute Physiology Score (SAPS), the Sequential Organ Failure Assessment Score (SOFA), which are used to gauge the severity of the disease, are linked to mortality in critically ill AKI patients but do not adequately predict their risk of dying due to lack of specificity [7].

Models such as MOSAIC, MALEDICT, and HEELENICC were developed based on patients undergoing continuous renal replacement therapy (CRRT). Recent meta-analyses have indicated that both intermittent and continuous hemodialysis result in comparable outcomes in terms of mortality and renal recovery. However, CRRT remains unaffordable for many low- and middle-income countries[8].

The aim of our study is development and validation of risk prediction model for in-hospital mortality of adult acute kidney injury patients requiring intermittent hemodialysis.

## 2. Materials and Methods

### 2.1 Source of data

The development cohort included 829 patients and retrospective follow up study was conducted from October 02, 2016 to May 18, 2025 and a data extraction tool was developed, and the entire populations were sampled and data were collected from patient’s medical chart on patient demographics, clinical parameters and laboratory values.

### 2.2 Participants

The study was conducted at the University of Gondar and Felege Hiwot Comprehensive Specialized Hospitals, two of the primary governmental healthcare institutions providing dialysis services in northwest Ethiopia. Inclusion criteria: age>18years, Base line creatinine ≤ 1.5 mg/dL, abdominal ultrasound finding of normal kidney size and no normocytic normochromic anemia, and patients required intermittent Hemodialysis. Exclusion criteria: Base line creatinine ≤ 1.5 mg/dL and patients with normocytic normochromic anemia, those required intermittent hemodialysis for dialyzable drug overdose and no stage 3AKI, AKI on CKD patients who required intermittent hemodialysis

### 2.3 Outcome

In-hospital mortality refers to the death of an adult patient diagnosed with acute kidney injury and initiated on intermittent hemodialysis, which occurs during the same hospitalization period and before discharge from the hospital.

### 2.4 Predictors

The following variables were extracted from the relevant literature and clinical records:

#### Demographic Characteristics

Sex, age and residence

#### Clinical Parameters

Mean arterial pressure, Urine output per hour, respiratory rate, GCS, DM, CVD, immunosuppression, ICU admission, use of vasopressors, Use of mechanical ventilation, post-surgery

#### The last laboratory test within 24 h prior to receiving IRRT

platelet, potassium, creatinine, BUN, WBC and hemoglobin.

#### Operational definition

##### CVD

is defined as the presence of one or more of the following clinically diagnosed conditions[9]

1. Heart failure of any etiology, confirmed by clinical assessment or imaging
2. Acute coronary syndrome (ACS), including unstable angina, non-ST elevation myocardial infarction (NSTEMI), or ST elevation myocardial infarction (STEMI) or
3. Rheumatic valvular heart disease (RVHD), confirmed by clinical diagnosis and/or echocardiographic evidence of valvular involvement due to rheumatic fever.

##### Immunosuppression

is defined as a condition in which a patient’s immune system is compromised due to one or more of the following:[10, 11]

1. a diagnosis of cancer (particularly hematologic malignancies or those undergoing chemotherapy or radiotherapy),
2. a diagnosis of Human Immunodeficiency Virus (HIV) infection
3. Current or recent use (within the last 3 months) of systemic corticosteroids

##### Baseline creatinine

Baseline kidney function defined as the most recent serum creatinine between 1 and 365 days prior to hospitalization [12].

### 2.5 Sample Size

The minimum sample size was calculated using the pmsampsize package in R software version 4.5.0, which assumes the following three default criteria [13]:

a. A minimum shrinkage of 0.9
b. The absolute difference between the apparent R^2^ and the adjusted R^2^ is set to be 0.05
c. The overall risk was estimated with a margin of error no greater than 0.05

**With a synthax**: n<-pmsampsize (type=“b”,nag rsquared = .28,parameters= 20, Incidence=0.29)(**table 1**)

**Table 1:**
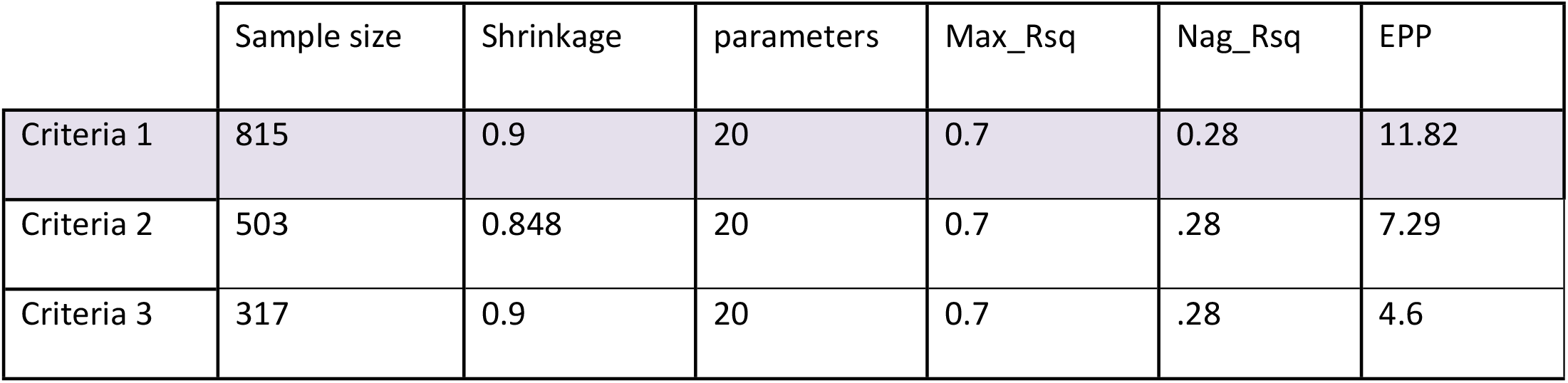
Pmsampsize criteria output.

|  | Sample size | Shrinkage | parameters | Max_Rsq | Nag_Rsq | EPP |
| --- | --- | --- | --- | --- | --- | --- |
| Criteria 1 | 815 | 0.9 | 20 | 0.7 | 0.28 | 11.82 |
| Criteria 2 | 503 | 0.848 | 20 | 0.7 | .28 | 7.29 |
| Criteria 3 | 317 | 0.9 | 20 | 0.7 | .28 | 4.6 |

### 2.6 Handling of Missing data

In the development dataset, there were missing data for few variables. Assumed that the data were missing at random and filled in missing data using a predictive mean matching method of multiple imputations, performed five multiple imputations with a maximum of fifty iterations via *mice* package in R software 4.5.0 version.

### 2.7 Statistical analysis

#### 2.7.1 Model Development

A Q-Q plot used to assess the normality of the continuous variables, and cubic spline functions were used to assess the linearity of the relationship. Continuous variables that did not conform to normal or linear distributions were transformed to log to address skewness and converted to categorical covariates based on their clinical significance.

Descriptive analysis was done using calculations of proportions for qualitative variables (frequencies and percentages) and means (normally distributed) and median (Skew) for quantitative variables.

Binary logistic regression was undertaken to select variables to predict the presence of the outcome, in-hospital mortality of acute kidney injury requiring intermittent hemodialysis. First, a bi-variable regression analysis was used to quantify the association of each potential determinant with in-hospital mortality of acute kidney injury that underwent intermittent hemodialysis using likelihood ratio test for inclusion in multivariable regression analysis. Variables with a p value<0.2 in the bi-variable analysis were included in the multivariable regression analysis. A stepwise backward elimination technique was used and the role of each predictor in the multivariable analysis was assessed by the Akaike *Information Criterion(AIC)*, with a stopping rule of p<0.20. Presence of multicollinearity was checked using variance inflation factor and a value greater than five indicates the presence of multicollinearity. A risk prediction model was developed from a reduced multivariable regression model, including significant variables at a p value of <0.05

Finally, Using the final model a nomogram was constructed that could provide clinicians with an intuitive and quantitative tool for predicting in-hospital mortality of AKI patients undergoing intermittent hemodialysis.

#### 2.7.2 Model Validation

Internal validation was performed with the enhanced bootstrap technique, in which regression models were fitted in1000 bootstrap replicates, drawn with replacement from the development cohort. The model was refitted in each bootstrap replicate and tested using the original sample to estimate optimism in the model performance.

#### 2.7.3 Model Performance evaluation

The model performance discrimination was evaluated with the area under the receiver operator characteristic curve (AUC).The model calibration was evaluated with Hosmer_Lemeshow test (P>0.05) and calibration plots. The overall performance of the model was assessed with brier score.Decision curve analysis (DCA) curves were constructed to assess the clinical usefulness of the model. Nomogram was constructed to assess the probability of each potential predictor for in-hospital mortality of acute kidney injury patients requiring intermittent hemodialysis.

### 2.8 Reporting

To ensure clarity, reproducibility, and completeness in presenting the model, TRIPOD checklist was followed.

### 2.9 Ethical Considerations

An official letter was obtained from the Department of Epidemiology and Biostatistics and subsequently provided by the Hospital Admission and Department of Internal Medicine for both university of Gondar and Felege Hiwot Comprehensive specialized Hospitals. The study was approved by the Institutional Review Board of the University of Gondar Comprehensive Specialized Hospital. Permission was sought from the institutions to use patient information for the study.

## 3. Result

### 3.1 Socio-demographic, Clinical and laboratory characteristics of study participants

#### 3.1.1 Socio-demographic characteristics of study participants

A total of 829 adult patients diagnosed with acute kidney injury (AKI) required intermittent hemodialysis (IHD) were included in the study among these, 582 patients (70.2%) survived until discharge, while 247 patients (29.8%) died during hospitalization. The median age of the study participants was 50.6 years; the median age of those who died was 64 years compared to 45 years for those who survived. Males constituted 57% of the study population **(table2**).

**Table 2:** Socio-demographic of adult acute kidney injury patients requiring intermittent hemodialysis at University of Gondar and FelegeHiwot Comprehensive Specialized Hospitals, 2025.

| Variables |  | Alive | Died | ALL |
| --- | --- | --- | --- | --- |
|  |  | 582 | 247 | 829 |
| Sex | Male, n (%) | 321(67.83) | 152(32.22) | 473(57) |
|  | Female, n (%) | 261(73.32) | 95(26.72) | 356 (43) |
| Residence | Rural, n (%) | 244(59) | 171(41) | 415(51) |
|  | Urban, n (%) | 338(82) | 76(18) | 414(49) |
| Age(median) |  | 45 | 64 | 50.62 |

#### 3.1.2 Clinical Parameters of the study participants

ICU admission was documented in 58.5% of the patients, and among those, 37.52 % were died. 76.20% of patients use mechanical ventilation, of this 30% were died and among 76.20% use of vasopressors 33% were deceased. Post-surgical causes of AKI were 15.20%, of this 11.70 % were died. Comorbid conditions such as among those having cardiovascular disease, 51% were deceased and among those having immunosuppression, 35% were died. The average mean arterial pressure and the Glasgow Coma Scale (GCS) scores were 84.74 and 10 in patients who died respectively (**table3**).

**Table 3:** Clinical Characteristics of adult acute kidney injury patients required intermittent hemodialysis at University of Gondar and FelegeHiwot Comprehensive Specialized Hospitals, 2025.

| Variables |  | Alive | Died | All |
| --- | --- | --- | --- | --- |
|  |  | 582 | 247 | 829 |
| ICU Admission | Yes, n (%) | 303(62.47) | 182(37.52) | 485(58.50) |
|  | No, n (%) | 279(81) | 65(19) | 344(41.50) |
| Use Of MV | Yes, n (%) | 442(70) | 190(30) | 632(76.20) |
|  | No, n (%) | 140(71) | 57(29) | 197(23.80) |
| Use Of Vasopressors | Yes, n (%) | 423(67) | 209(33) | 632(76.20) |
|  | No, n (%) | 159(81) | 38(19) | 197(23.80) |
| Post-Surgery | Yes, n (%) | 29(23) | 97(77) | 126(15.20) |
|  | No, n (%) | 553(78.74) | 150(21.32) | 703(84.80) |
| Immunosuppression | Yes, n (%) | 424(65) | 228(35) | 652(78.60) |
|  | No, n (%) | 158(89) | 19(11) | 177(21.40) |
| CVD | Yes, n (%) | 185(49) | 190(51) | 375(45.20) |
|  | No, n (%) | 397(87) | 57(13) | 454(54.80) |
| DM | Yes, n (%) | 148(56.5) | 114(43.50) | 262(31.60) |
|  | No, n (%) | 434(76.5) | 133(23.50) | 567(68.40) |
| Urine output/hr/kg ( in ml) (mean) |  | 1.10 | 0.90 | 1.029 |
| MAP mmHg(mean) |  | 82 | 92 | 84.74 |
| GCS(mean) |  | 11.60 | 6.30 | 10.10 |
| RR bpm(mean) |  | 19 | 20 | 19.48 |

##### Laboratory Parameters of the study participants

In the laboratory profile, non-survivors had a mean value of 11,330/microliter of white blood cell counts, and median values of serum creatinine and platelet counts were 8.92mg/dl and mean 75,000/microliter respectively. The mean values of blood urea nitrogen and potassium were 76mg/dl and 5.8meq/l among non-survivors respectively, and mean hemoglobin value of 12.8mg/dl among deceased (**table4**).

**Table 4:** Laboratory values of adult acute kidney injury patients requiring intermittent hemodialysis at University of Gondar and FelegeHiwot Comprehensive Specialized Hospitals, 2025.

| Variables | Alive | Died | All |
| --- | --- | --- | --- |
|  | 582 | 247 | 829 |
| Plt($10^3$/microliter)(median) | 180 | 75 | 147.43 |
| Cr(mg/dl)(median) | 65.12 | 8.92 | 6.25 |
| BUN (mg/dl)(mean) | 52.4 | 76 | 55.7 |
| K<sup>+</sup>meq/L(mean) | 4.2 | 5.8 | 3.98 |
| WBC$10^3$/microliter (mean) | 7.9 | 11.3 | 8.5 |
| Hgb(mean)(mg/dl) | 12.6 | 12.8 | 12.65 |

### 3.2 Model development

A stepwise backward elimination technique was used and the role of each predictor in the multivariable analysis was assessed by the Akaike *Information Criterion(AIC)*, with a stopping rule of p<0.20. A risk prediction model was developed from a reduced multivariable regression model, including significant variables at a p value of <0.05.The VIFs of the screened variables were all <5. Seven variables (age, vasopressor use, immunosuppression, platelet count, Serum creatinine, CVD and Use of MV) were finally included in the model, which was used to plot the nomogram (**table5**).

**Table 5:** Potential predictors included in the final reduced model for multivariable risk prediction model for adult acute kidney injury patients requiring intermittent hemodialysis at UGCSH and FHCSH, Northwest Ethiopia, 2025.

| Variables |  | COR(95%CI) | AOR(95% CI) | p_value |
| --- | --- | --- | --- | --- |
| Age | 15-24 | 1 | 1 |  |
|  | 25-44 | 1.12(1.1-1.17) | 1.04(1.02-1.06) | 0.048 |
|  | 45-54 | 1.32(1.28-1.34) | 1.32(1.28-1.34) | .043 |
|  | 60-74 | 2.11(1.98-2.33) | 1.90(1.87-2.10) | 0.025 |
|  | ≥ 75 | 2.55(2.49-2.57) | 2.03(1.98-2.11) | .014 |
| Use of Mechanical Ventilation | No | 1 | 1 |  |
|  | Yes | 2.0(1.85-1.91) | 1.8(1.74-1.83) | .001 |
| Use of vasopressors | No | 1 | 1 |  |
|  | Yes | 2.32(2.28-2.35) | 2.25(2.2-2.3) | .015 |
| Immunosuppression | No | 1 | 1 |  |
|  | Yes | 3.3(3.27-3.32) | 3.3 (3.25-3.33) | .002 |
| CVD | No | 1 | 1 |  |
|  | Yes | 2.71(2.68-2.74) | 2.6(2.51-2.7) | .018 |
| Platelet (10<sup>3</sup>/microliter) | >150 | 1 | 1 |  |
|  | 100-149 | 1.20 (1.15-1.23) | 1.13(1.11-1.15) | 0.032 |
|  | 50-99 | 1.80(1.77-1.82) | 1.61(1.57-1.63) | 0.027 |
|  | <50 | 3.20 (3.18-3.22) | 1.90(1.88-1.92) | 0.01 |
| Serum creatinine (mg/dl) | 0.5-1.49 | 1 | 1 |  |
|  | 1.5-2.0 | 1.3(1.29-1.33) | 1.2(1.20-1.24) | 0.044 |
|  | >2.0 | 1.5(1.48-1.54) | 1.3(1.27-1.32) | .012 |
|  | 25-44 | 1.12(1.1-1.17) | 1.04(1.02-1.06) | 0.048 |
|  | 45-54 | 1.32(1.28-1.34) | 1.32(1.28-1.34) | .043 |

**Note:** 1-

### 3.3 Model performance

The developed model demonstrated a discriminatory power, with an area under the curve (AUC) of 0.85(95%CI, 0.8-0.89) in the development dataset (**fig2A**) and 0.8(95%CI, 0.76-0.83) after internal validation using bootstrap resampling (**fig2B**). The calibration of the model, which is the agreement between predicted and observed mortality probabilities assessed via Hosmer_Lemshow test, p=0.35 and visualized using calibration plot (**fig2C**) after bootstrapping. The overall performance was assessed via a brier score=.06 and 0.171 for development and validation datasets respectively. The confusion matrix **(table6**) and the performance measured values were listed below (**table7**).

**Fig 1:**
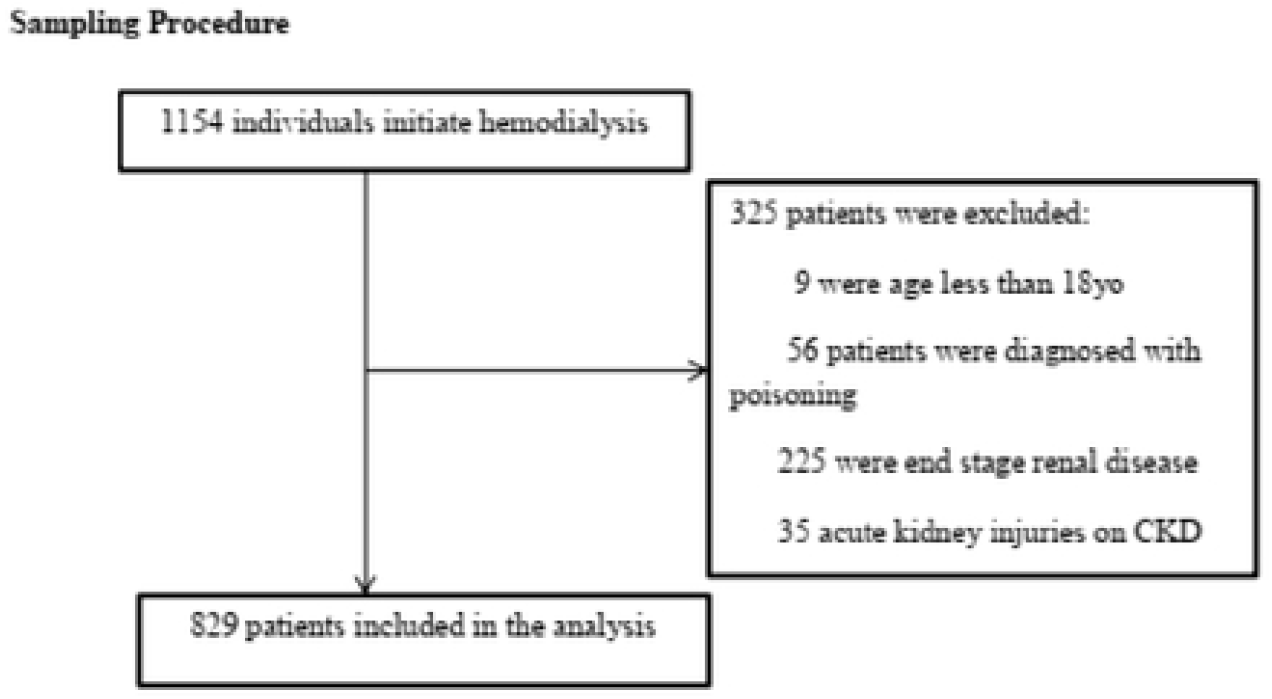
Sampling Procedure. Sampling procedure for in-hospital mortality of acute kidney injury requiring intermittent hemodialysis

**Fig 2:**
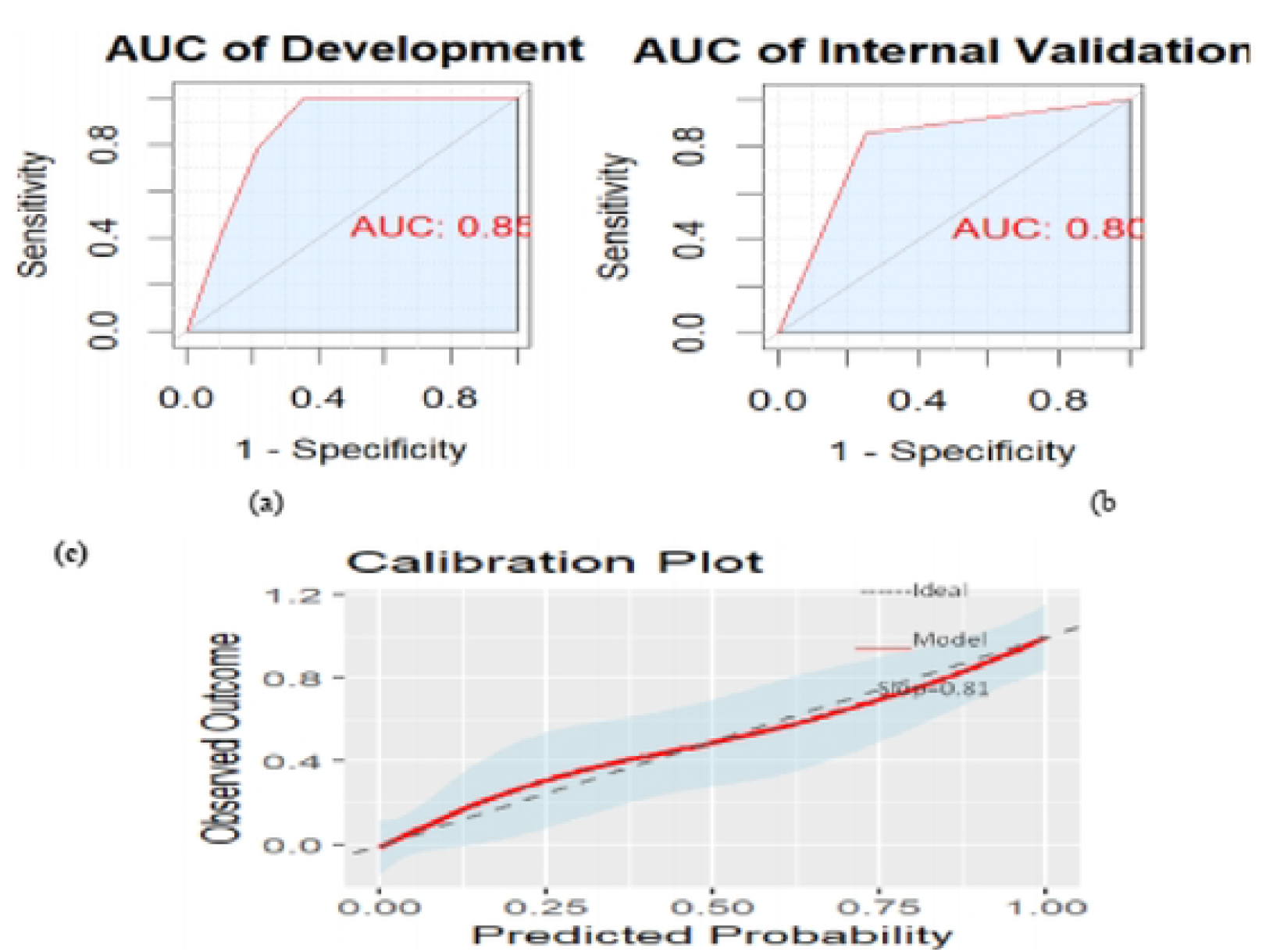
(A) AUC of a development dataset model (B) AUC of internal validation dataset after bootstrap resampling (C) Calibration plot with smooth-loess of internal validation

**Table 6:**
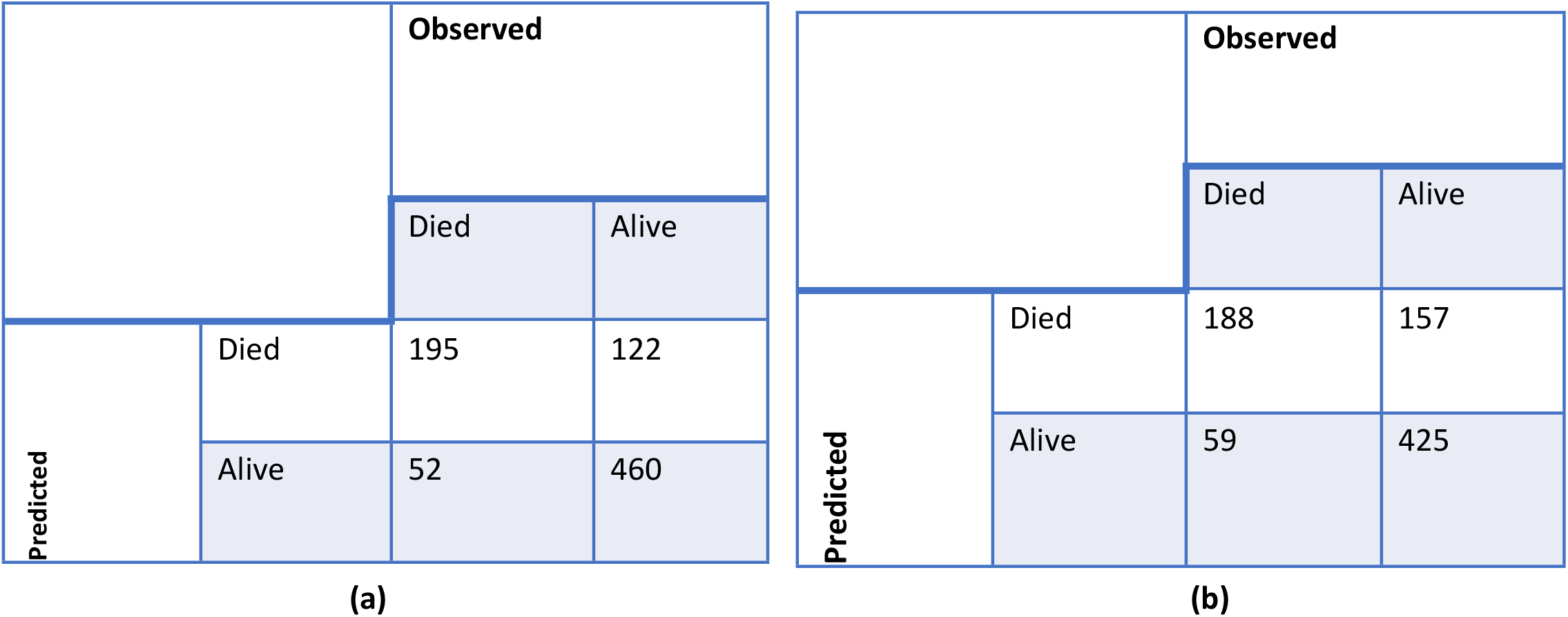
Confusion matrix for predicting in-hospital mortality of adult AKI patients required intermittent hemodialysis **(a)** development dataset **(b)** after bootstrapping.

**Table 7:** Model performance values of adult acute kidney injury patients required intermittent hemodialysis.

| Model performance measures | Development Model | Internal Validation model |
| --- | --- | --- |
| AUC | 0.85 | 0.8 |
| Sensitivity | 0.79 | 0.76 |
| Specificity | 0.79 | 0.73 |
| Positive Predictive Value(precision) | 0.62 | 0.55 |
| Negative predictive value | 0.9 | 0.88 |
| Brier Score | 0.06 | 0.171 |
| Hosmer_Lemeshow | 0.59 | 0.35 |
| Accuracy | 0.79 | 0.74 |

### 3.4 Clinical usefulness and nomogram

The DCA shows the net benefit of the model at different ranges of risk threshold probabilities which help in making treatment decision. (**fig3A**).Nomogram estimates the individual patient’s probability of in hospital mortality of adult acute kidney injury patients required intermittent hemodialysis. (**fig3B**).

**Fig 3:**
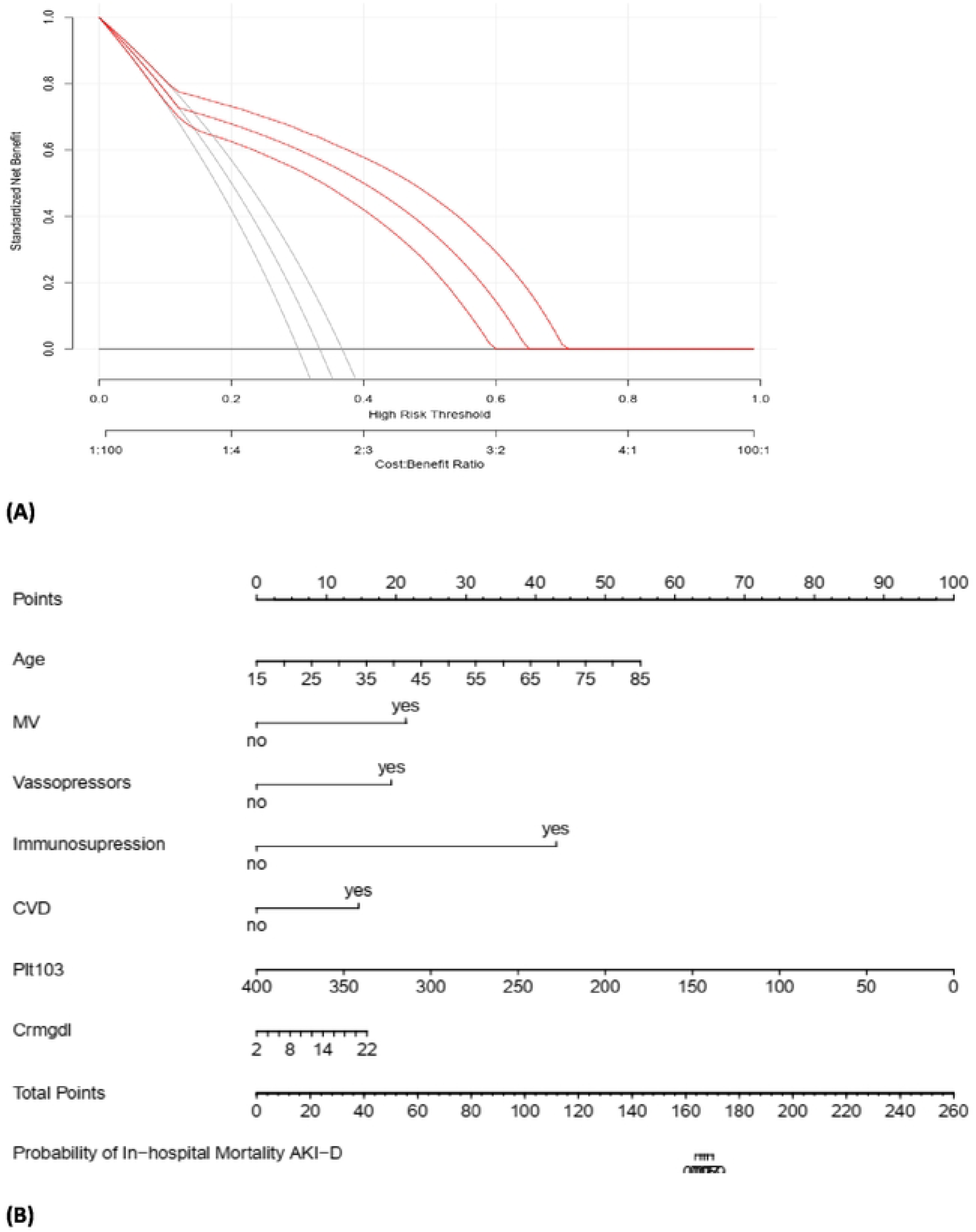
(A) Decision curve analysis showing the clinical utility of the model (B) Nomogram: estimates the individual probability

## 4. Discussion

This study developed and internally validated a risk prediction model for in-hospital mortality among adult patients with acute kidney injury requiring intermittent hemodialysis in two comprehensive specialized hospitals in Northwest Ethiopia. The model incorporated a combination of clinical, demographic, and laboratory parameters and demonstrated a good model performance, as evidenced by an area under the receiver operating characteristic curve (AUC) of 0.80, indicating good discriminative ability[14].

The Hosmer–Lemeshow goodness-of-fit test yielded a p-value of 0.35, suggesting that there is no significant difference between the observed and predicted probabilities[15]. The Brier Score was 0.06, indicating a good overall performance of the model[15, 16].

The overall in-hospital mortality in this study was 29.8%, which is consistent with findings from similar settings, including Ethiopia (29.1%)[17] and other sub-Saharan African countries like the Democratic Republic of Congo (25.1%) and South Africa (31%)[18]. But remains lower than some global figures which report mortality rates as high as 49% among AKI-D patients[17]. Regional differences in case severity, resource availability, and dialysis initiation time may be the cause of this disparity [19].

Among non-survivors, the usage of vasopressors and mechanical ventilation was significantly greater. The deceased had a higher prevalence of post-surgical causes of AKI and concomitant comorbidities such immunosuppression and cardiovascular disease. The Glasgow Coma Scale (GCS) scores and mean arterial pressure were also significantly lower in the deceased patients.

In the laboratory profile, non-survivors had higher white blood cell counts and serum creatinine levels and lower platelet counts. Bi-variable analysis showed platelet count, BUN, serum creatinine and serum potassium were significantly associated with mortality.

The model’s performance, evidenced by an AUC of 0.8 internal validations, is better than existing risk models developed in other settings. For instance, the MALEDICT model had an AUC of 0.77, and the MOSIAC model, which reported an AUC of 0.772[20, 21].

The Mosaic model itself has been shown to outperform traditional prognostic tools such as SOFA and APACHE II [21]. The HELENICC model which was yet not externally validated and had an AUC of 0.82 for internal validation, specifically developed for predicting seven-day mortality in patients with septic acute kidney injury (AKI) requiring hemodialysis [22]. Decision curve analysis (DCA) was employed to assess the clinical net benefit of our model across a range of threshold probabilities. The DCA curve demonstrated that the model offers greater net benefit than either treating all or treating none of the patients, particularly within the clinically relevant probability thresholds. This indicates that using the model to guide decisions can improve patient outcomes without unnecessary overtreatment[23].

To facilitate clinical application of this model, a nomogram incorporating the identified independent predictors was constructed. The nomogram provides an individualized estimation of in-hospital mortality risk in AKI patients undergoing intermittent hemodialysis, translating complex regression outputs into an accessible graphical tool for bedside decision-making. Its visual format supports shared decision-making between clinicians and patients or their families at the time of initiating renal replacement therapy [23-25].

### Strength and limitation of the study

First, the study is multicenter and assessed different predictors for in-hospital mortality among adult patients with acute kidney injury requiring intermittent hemodialysis. Second, decision curve analysis and the development of a nomogram enhanced the practical utility of the model, supporting its potential for bedside clinical decision-making.

Despite these strengths, the study has limitations. First, some potentially useful predictors, such as serum lactate, serum bilirubin were not available in the dataset due to unavailability of data. Second, External validation was not performed, which may affect generalizability. And finally, the findings of this study may not represent individuals under the age of 18years.

## 5. Conclusion

This study successfully developed and internally validated a risk prediction model for mortality in adult patients with acute kidney injury requiring intermittent hemodialysis. The model demonstrates good potential to support clinical decision-making by enabling individualized mortality risk estimation at the time of initiating renal replacement therapy. With external validation in independent cohorts, it could become a valuable tool to guide prognosis and facilitate shared decision-making in clinical practice.

## Data Availability

there is data availability as required

## 6. Acknowledgments

I would like to express my sincere gratitude to the University of Gondar, Department of Epidemiology and Biostatistics, for granting me this incredible opportunity. My heartfelt appreciation also goes to my beloveds, Mr. Dessie Abebaw and Mr. Lakew Asmare, for their invaluable comments and unwavering support. I extend my thanks to both the University of Gondar and Felege Hiwot Comprehensive Specialized Hospitals dialysis unit team leaders for providing the preliminary information necessary for obtaining data. I would also like to thank all contributors and participants who make this thesis report possible.

## 7. Authors’ contribution

**AW**: Formal analysis; Visualization; writing-original draft; Writing-review and editing

**DA:** Conceptualization; Methodology

**LA:** Software; Validation

## 10. Spportive information captions

**S1 fig:** Q_Q plot showing normality of continous variables (A) platelet count (B) Potassium level (C) serum creatinine

**S2 image**: Ethical clearance

